# Prevalence and determinants of Kaposi’s sarcoma-associated herpesvirus (KSHV) antibody positivity among adults living with HIV in East Africa

**DOI:** 10.1101/2022.12.20.22283702

**Authors:** Chandana K. Gowdara, Helen Byakwaga, Sheila C. Dollard, Conrad K. Muzoora, David V. Glidden, Peter W. Hunt, Bosco M. Bwana, Jessica E. Haberer, David R. Bangsberg, Jeffrey N. Martin

**Affiliations:** University of California, San Francisco; Mbarara University of Science and Technology, Mbarara, Uganda; Centers for Disease Control and Prevention, Atlanta, Georgia; Massachusetts General Hospital, Boston, Massachusetts; Oregon Health and Science University-Portland State University School of Public Health, Portland, Oregon

**Keywords:** Kaposi’s sarcoma-associated herpesvirus, antibodies, seroprevalence, HIV, determinants, Uganda, East Africa, Africa

## Abstract

**Background:** Persons living with HIV (PLHIV) who are also infected with Kaposi sarcoma-associated herpesvirus (KSHV) constitute a group among the highest risk for Kaposi sarcoma (KS). As such, understanding KSHV prevalence amongst PLHIV is important for the control of KS. To date, data on KSHV prevalence amongst PLHIV in East Africa — one of the world’s hotbeds for KS — is both sparse and variable.

**Methods:** In a cross-sectional design, we studied consecutive adult PLHIV identified just prior to starting antiretroviral therapy at an ambulatory HIV clinic in Mbarara, Uganda. Results from two enzyme immunoassays (with synthetic K8.1 and ORF 65 antigens as targets) and one immunofluorescence assay (using induced BCBL cells) to detect antibodies to KSHV were combined to classify KSHV antibody positivity.

**Results:** We evaluated 727 PLHIV between 2005 to 2013; median age was 34 years (interquartile range (IQR): 28-40), 69% were women, and median CD4 count was 167 cells/µl (IQR: 95-260). Prevalence of KSHV antibody positivity was 42% (95% CI: 38%-46%), with little substantive change after several correction approaches, including Rogan-Gladen. Adjusted prevalence of KSHV antibody positivity was 1.6 times (95% CI: 1.3-1.9) higher in men than women; adjusted absolute prevalence difference was +0.20 (95% CI: +0.11 to +0.30). Lack of formal education (prevalence ratio=1.6 comparing no school to ≥ 4 years of secondary school; 95% CI: 1.1-2.3) was also associated with KSHV infection. We found no strong evidence for a role for age, alcohol use, or other measurements of sexual behavior, SES, or well-being in the occurrence of KSHV antibody positivity.

**Conclusion:** Among adult PLHIV in western Uganda, KSHV prevalence is estimated at 42%, with little change after several approaches to correction for antibody detection inaccuracy. This estimate differs from several others in the region (up to 83%), highlighting need for inter-assay comparison studies using identical local specimens. To the extent HIV does not influence KSHV acquisition, the findings may also represent KSHV prevalence in the general population. The large-magnitude effect of sex and education on KSHV acquisition motivates an accelerated search for mechanisms. The sex effect, in part, may explain the higher incidence of KS among men.

---

Even before the AIDS epidemic, portions of East Africa had amongst the highest incidences of Kaposi sarcoma (KS) in the world.^1^ Widespread HIV infection in the region accelerated^2-4^ this already high incidence and transformed KS into a much more aggressive condition with poorer survival.^5, 6^ The advent of HIV-related KS also speeded the search for the long sought-after infectious agent of KS,^7^ culminating in the discovery of KSHV.^8^ Soon thereafter, it became recognized that KSHV is necessary but not sufficient for development of KS,^9, 10^ and HIV infection is one of the strongest complementary co-factors.^7, 11^ As such, persons living with HIV infection (PLHIV) form a group among highest risk for KS if they are coinfected with KSHV.^12^ Accordingly, knowledge of KSHV prevalence as well as determinants of KSHV infection among PLHIV represent critical yardsticks to understand the threat of KS in communities and factors to target in an attempt to mitigate its occurrence.

Initial research on KSHV prevalence in PLHIV in East Africa has featured extensive variability in terms of persons evaluated^13-19^ and assays^13, 20, 21^ used to document KSHV antibody positivity. Findings have also been variable, with prevalence estimates ranging between 29% in the rural Rakai district of Uganda^22^ and 83% in the Kalungu district of Uganda.^14^ Regarding determinants of KSHV infection among PLHIV, very little work has been performed in East Africa, with the closest analogous research emanating from other parts of Africa.^23, 24^

To better understand the prevalence and determinants of KSHV infection among PLHIV in East Africa, we focused on PLHIV identified just prior to starting antiretroviral therapy (ART) in a prototypic municipal HIV clinic caring for a diverse spectrum of adults in Uganda. We took advantage of a large and well-characterized research cohort, previously assembled to study clinical course following initiation of ART. In addition, to the extent that HIV infection per se does not influence KSHV prevalence,^25-27^ we also argue that our estimate of KSHV seroprevalence made in this representative sample of PLHIV provides insights on KSHV seroprevalence in the general adult population in this region.

## Methods

### Overall Design

Using a cross-sectional design, we determined the prevalence and determinants of KSHV antibody positivity among adult PLHIV in Uganda just prior to their initiation of ART.

### Participants

Participants were all those with a baseline visit in the Uganda AIDS Rural Treatment Outcomes (UARTO) research cohort. UARTO was a consecutive sample of consenting ambulatory adult PLHIV (18 years or older) initiating ART at and residing within 60 kilometers of the Immune Suppression Syndrome (ISS) Clinic of the Mbarara Regional Referral Hospital in southwestern Uganda. While the clinic is located in the urban setting of Mbarara (population estimated at 82,000 in 2010), the patients cared for are derived from a wide catchment of urban, peri-urban and rural residences. Patients were deemed eligible to begin ART by local clinic providers, not affiliated with the research cohort, according to clinical indications following Ugandan Ministry of Health and WHO guidelines. In the first wave of enrollment into UARTO, patients were recruited between June 2005 and November 2008, and a second wave of enrollment occurred between June 2011 and July 2013. These periods featured CD4+ T cell- and other clinical indication-derived thresholds for ART initiation; this was prior to the current “Treat-All” era. All participants provided written informed consent.

### Measurements

#### Questionnaire-based

Participants completed standardized interviewer-administered surveys detailing demographic characteristics, asset holdings (via the Filmer-Pritchett asset index^28^), frequency of hunger, HIV-related clinical history, health-related quality of life (via the Medical Outcome Study-HIV^29^), alcohol use (via the Alcohol Use Disorder Identification Test^30^), and sexual behavior.

#### Laboratory-based

Venous blood samples were drawn from participants prior to initiation of ART. Two enzyme immunoassays (EIA) (with synthetic K8.1 and ORF 65 antigens as target) and one indirect immunofluorescence assay (IFA) (using an induced KSHV-infected BCBL cell line expressing lytic-phase KSHV antigens) were performed at the Centers for Disease Control and Prevention to detect antibodies to KSHV.^31-34^ The findings from these three assays were interpreted according to a previously described algorithm, leading to a final overall result.^35^ In this algorithm, specimens reactive in any two tests or the immunofluorescence assay alone were classified as KSHV-antibody-positive. This algorithm has an estimated specificity of 97.5% for the diagnosis of KSHV infection, as evidenced by testing blood donors in the U.S.,^36^ and sensitivity of 96.3% based on testing patients with KS.^37^ Plasma HIV RNA level was measured using the Amplicor® HIV Monitor version 1.5 or the COBAS® TaqMan HIV-1 version 1.0 assays (Roche, Branchburg, NJ). CD4+ T cell counts were assessed using the FACSCalibur™ system (Becton Dickinson, San Jose, CA).

#### Literature search

To identify all published literature regarding KSHV seroprevalence in East Africa, we searched Medline (PubMed), EMBASE, SCOPUS and Web of Science from 1998–2022, according to the PRISMA guidelines, for all articles providing data on the prevalence of KSHV infection by antibody testing in Djibouti, Eritrea, Ethiopia, Kenya, Somalia, Tanzania, and Uganda. The search strategies are listed in the Supplementary Material.

### Statistical Analysis

Because the measurement of KSHV antibodies is imperfect, we calculated, in addition to the base KSHV antibody positivity derived by the combined assay algorithm, a range of corrected antibody positivity values using the Rogan-Gladen approach, which takes into account presumed assay sensitivity and specificity imperfections.^38, 39^ We also determined overall KSHV seroprevalence using a rule in which reactivity on any individual assay equates to overall antibody positivity.

To evaluate determinants of KSHV antibody positivity, we used antibody status derived from the combination assay algorithm. We focused on three constructs as the primary proximal exposures of interest: a) non-sexual horizontal contact with other humans in childhood; b) non-sexual horizontal contact in adulthood; and c) sexual contact in adulthood. For the first two constructs, we did not have direct measurements and instead have measurements that are upstream causes of the underlying constructs but may not be highly associated with the underlying construct. For the third construct (sexual contact in adulthood), we have several direct measurements. Our conceptualization of the system was depicted in a directed acyclic graph (DAG; Figure 1). Attempts to identify the causal effects of the various exposures of interest, including which variables to control for to remove confounding, was informed by the DAG.^40, 41^ This process resulted in several different regression models, each focusing on a primary exposure of interest, rather than a single final model.^42^ Relationships between exposure variables and outcome were estimated in a causal framework with prevalence ratios (PR) and prevalence differences, which were obtained with the margins command in Stata following logistic regression.^43^ This command uses parametric G-computation to average across counterfactual contrasts formed for each participant in the study population.^44^ In the regression models, continuous exposure variables were specified as restricted cubic splines. When a continuous variable was the primary exposure of interest, our measures of association contrasted participants who had values at the 75^th^ percentile of the distribution to participants with values at the 25^th^ percentile. The regression models were assessed for influential observations by comparing the coefficient value when an observation is included versus the coefficient when the same observation is excluded.^45^ For goodness-of-fit, the Hosmer-Lemeshow test was used.^46^ All analyses were performed in Stata (version 17; Stata Corporation, College Station, Texas).

**Figure 1.**
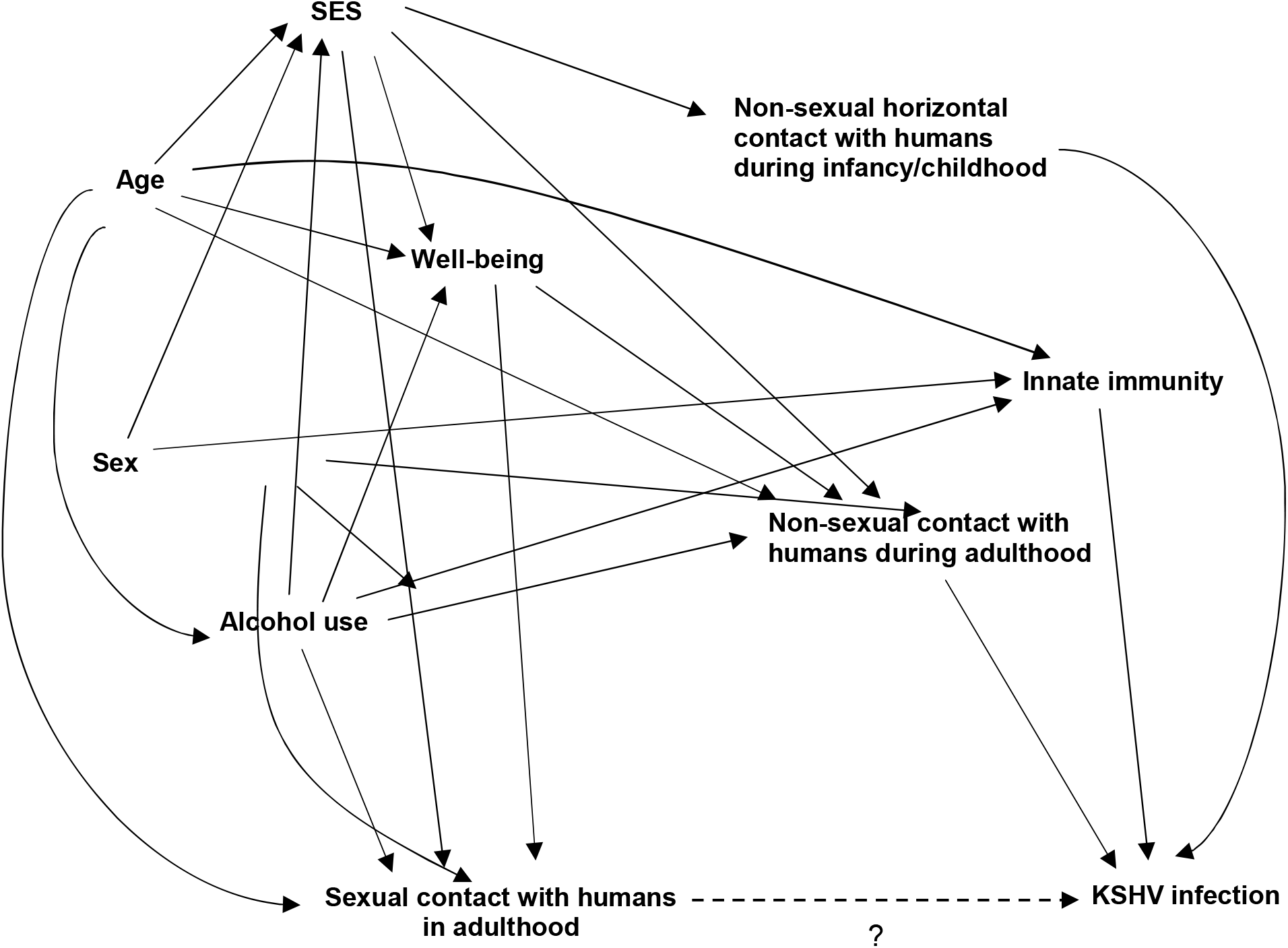
A directed acyclic graph (DAG) depicting our hypothesized conception of the system under investigation. We seek to estimate the causal contribution of age, biologic sex, alcohol use, socioeconomic status (SES; proxy measured by education, asset index, and frequency of hunger), well-being (proxy measured by a mental health quality-of-life index, physical health quality-of-life index; peripheral CD4+ T cell count; and history of major clinical co-morbidity in the form of tuberculosis and cryptococcosis), and sexual contact with humans during adulthood (measured by age of sexual debut, lifetime number of penile-vaginal sexual partners, circumcision history, and number of prior 3 month penile-vaginal partners) on the occurrence of KSHV infection (as measured by KSHV antibody positivity) in HIV-infected adults in Uganda just prior to initiation of antiretroviral therapy. The DAG was used to guide which variables to control for when assessing the independent contribution of the various constructs. In this rendition of the DAG, as depicted by the “?”, we are focusing on investigating the relationship between sexual contact with other humans in adulthood and KSHV infection. Other perspectives on this DAG can be used to investigate potential causal roles for the other variables (e.g., alcohol use).

## Results

### Characteristics of the Study Population

To investigate the prevalence and determinants of KSHV antibody positivity, we evaluated 727 ambulatory adult PLHIV initiating ART in Uganda. Median age was 34 years (interquartile range (IQR): 28 to 40), 69% were women, and 92% were ever married (Table 1). A median monthly income of $14.60 U.S. dollars (IQR: $5.90 to $41) and high frequency of hunger were evidence of the impoverished context of the region. The median CD4+ T cell count was 167 cells/µl (IQR: 95 to 260), and 100% of population had detectable plasma HIV RNA.

**Table 1.**
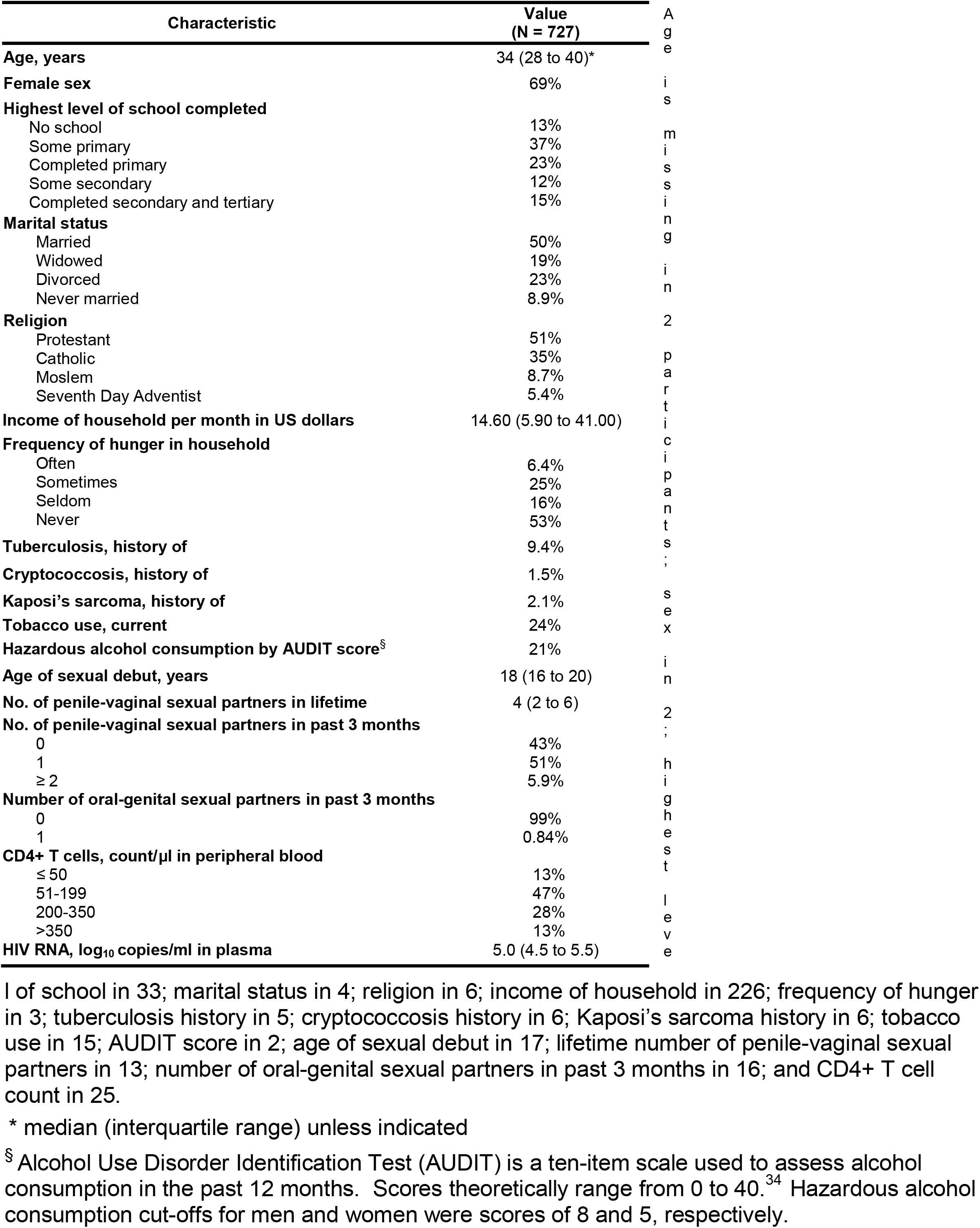
Characteristics of HIV-infected adults in southwestern Uganda just prior to initiation of antiretroviral therapy.

### Prevalence of KSHV Antibody Positivity

When evaluating the three antibody assays in isolation, reactivity was higher in the IFA (41%) and ORF K8.1 EIA (40%) compared to the ORF 65 EIA (25%) (p-value <0.001 for both using McNemar’s test) (Table 2). Although both yielded approximately 40% reactivity, the IFA and ORF K8.1 EIA were not fully concordant. A total of 247 (34%) participants were reactive on both assays, 364 (50%) were non-reactive on both; 38 (5.3%) reactive on the IFA but non-reactive on the ORF K8.1; 39 (5.4%) were reactive on the ORF K8.1 but not IFA; and 39 (5.4%) had a variety of combinations including an indeterminate value on one of the two assays. When evaluated in the laboratory’s previously established algorithm that combines results across all three assays, overall KSHV antibody positivity was 303/727 (42%; 95% CI: 38% to 45%). Among these 303 participants, KSHV antibody positivity was achieved by reactivity on all three assays in 142 (47%), reactivity in the IFA and one of the two EIAs in 112 (37%), reactivity in the IFA alone in 43 (14%), and reactivity in the two EIAs but not in the IFA in 6 (2%).

**Table 2.** Reactivity on three individual assays for detection of antibodies to KSHV (A) and patterns of reactivity (B) among HIV-infected adults in southwestern Uganda just prior to initiation of antiretroviral therapy.

| **Assay and Reactivity** | **Number (%)**<br>**(N=727)** |
| --- | --- |
| **EIA<sup>†</sup>: ORF K8.1** |  |
| Reactive | 289 (40%) |
| Non-reactive | 415 (57%) |
| Indeterminate | 23 (3.2%) |
| **EIA: ORF 65** |  |
| Reactive | 181 (25%) |
| Non-reactive | 527 (72%) |
| Indeterminate | 19 (2.6%) |
| **IFA<sup>‡</sup>** |  |
| Reactive | 297 (41%) |
| Non-reactive | 414 (57%) |
| Indeterminate | 16 (2.2%) |

**IFA = Reactive**
|  |  | **ORF 65** |  |  |
| --- | --- | --- | --- | --- |
|  |  | **Reactive** | **Non-reactive** | **Indeterminate** |
| **ORF K8.1** | **Reactive** | 142 (20%) | 98 (13%) | 7 (0.96%) |
|  | **Non-reactive** | 4 (0.55%) | 31 (4.3%) | 3 (0.41%) |
|  | **Indeterminate** | 3 (0.41%) | 8 (1.1%) | 1 (0.14%) |

**IFA = Non-reactive**

|  |  | **ORF 65** |  |  |
| --- | --- | --- | --- | --- |
|  |  | **Reactive** | **Non-reactive** | **Indeterminate** |
| **ORF K8.1** | **Reactive** | 3 (0.41%) | 35 (4.8%) | 1 (0.14%) |
|  | **Non-reactive** | 23 (3.2%) | 336 (46%) | 5 (0.69%) |
|  | **Indeterminate** | 3 (0.41%) | 6 (0.83%) | 2 (0.28%) |

**IFA = Indeterminate**

|  |  | **ORF 65** |  |  |
| --- | --- | --- | --- | --- |
|  |  | **Reactive** | **Non-reactive** | **Indeterminate** |
| **ORF K8.1** | **Reactive** | 3 (0.41%) | 0 | 0 |
|  | **Non-reactive** | 0 | 13 (1.8%) | 0 |
|  | **Indeterminate** | 0 | 0 | 0 |

<sup>†</sup> EIA denotes enzyme immunoassay
<sup>‡</sup> IFA denotes indirect immunofluorescence assay

Because the current state of the field in KSHV antibody detection does not feature a comprehensive spectrum of true positive and true negative gold standards, all estimated values of sensitivity and specificity from individual assays or combined assays/algorithms cannot be considered definitive. To address this, we used the approach of Rogan-Gladen to estimate the expected prevalence of KSHV antibody positivity after correcting for a range of assay sensitivity and specificity imperfections. Using the previously estimated 96.3% sensitivity and 97.5% specificity for the combined algorithm, the Rogan-Gladen correction was nearly unchanged, compared to the raw prevalence, at 42% (95% CI: 38% to 46%). When we concurrently varied estimates of sensitivity and specificity from 85% to 100%, we derived a range of corrected estimates from 31% (95% CI: 24% to 38%) to 49% (95% CI: 43% to 55%) (Figure 2). Finally, using a rule in which reactivity on any individual assay equates to overall antibody positivity, 371 (51%) were KSHV-antibody-positive.

**Figure 2.**
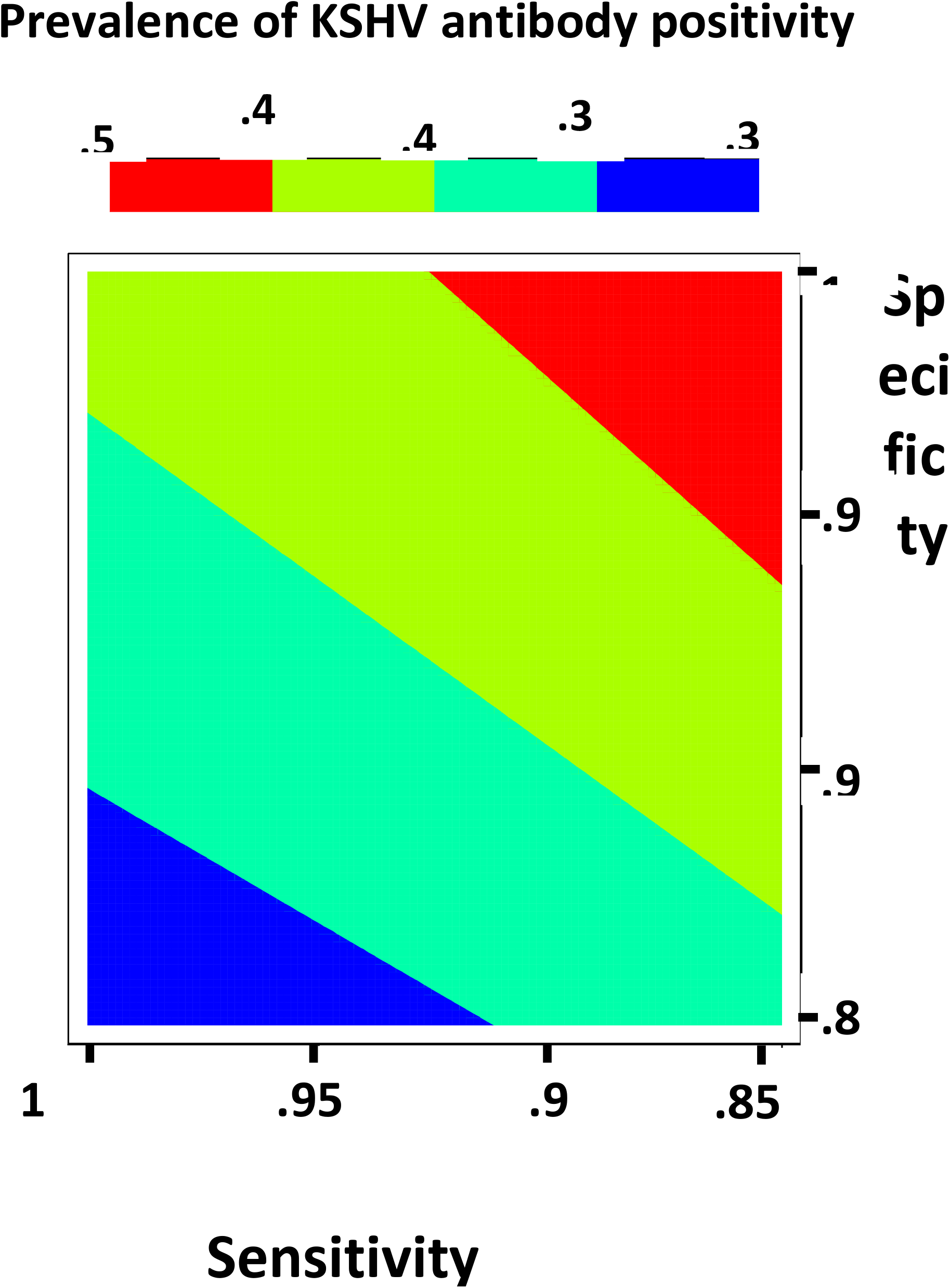
Contour plot depicting the range of corrected estimates, according to sensitivity and specificity of the testing algorithm, of the prevalence of KSHV antibody positivity among HIV-infected adults in southwestern Uganda just prior to initiation of antiretroviral therapy. The corrected estimates are calculated using the Rogan-Gladen formula, which combines the observed prevalence (0.42) and an assumed sensitivity and specificity of the testing algorithm to obtain the true prevalence, sampling error notwithstanding.

### Determinants of KSHV Antibody Positivity

Informed by our DAG, we evaluated several potential causal determinants of KSHV antibody positivity. Biological sex and education exhibited the largest magnitudes of effect (Table 3). After adjustment for several factors mediating indirect paths, the prevalence of KSHV antibody positivity in men was 1.6 times (95% CI: 1.3 to 1.9) higher than in women. This equates to an absolute adjusted prevalence difference of +0.20 (95% CI: +0.11 to +0.30). Lack of formal education (PR=1.6, comparing no school to ≥ 4 years of secondary school; 95% CI: 1.1-2.3, p=0.004 for trend) was associated with higher KSHV infection, equating to a prevalence difference of +0.17 (95% CI: +0.040 to +0.31). Older age at sexual debut (PR = 1.2, comparing 20 years to 16 years; 95% CI: 0.99 to1.3) and higher physical health score (a quality-of-life metric from the Medical Outcomes Study; PR = 1.3 comparing scores at the 75th to 25th percentile; 95% CI: 1.03 to 1.5) were also independently associated with higher KSHV prevalence but at lower magnitude. We found no strong evidence for a role for age, alcohol use, or other measurements of sexual behavior, socioeconomic status, or well-being in the occurrence of KSHV antibody positivity.

**Table 3.**
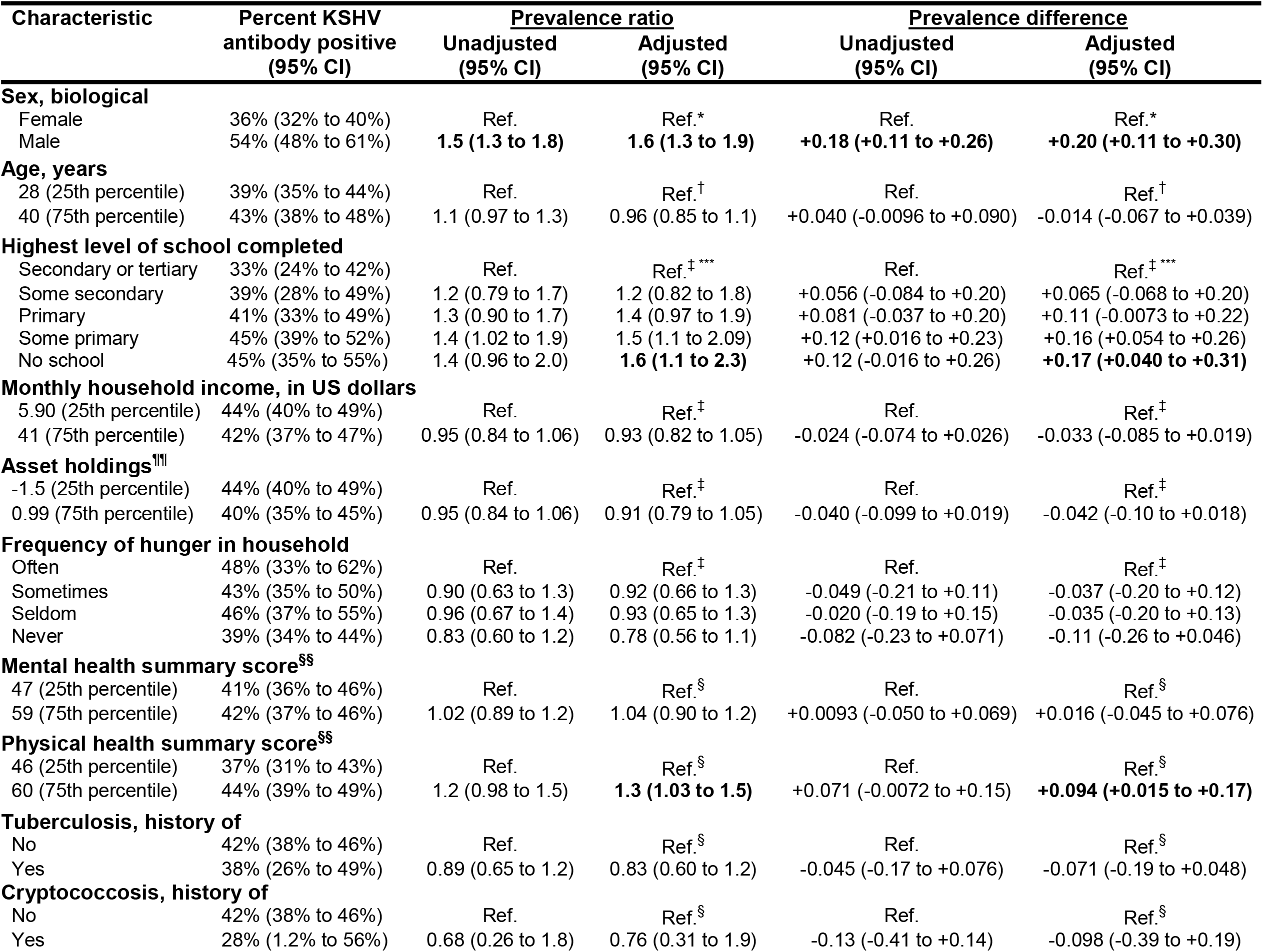
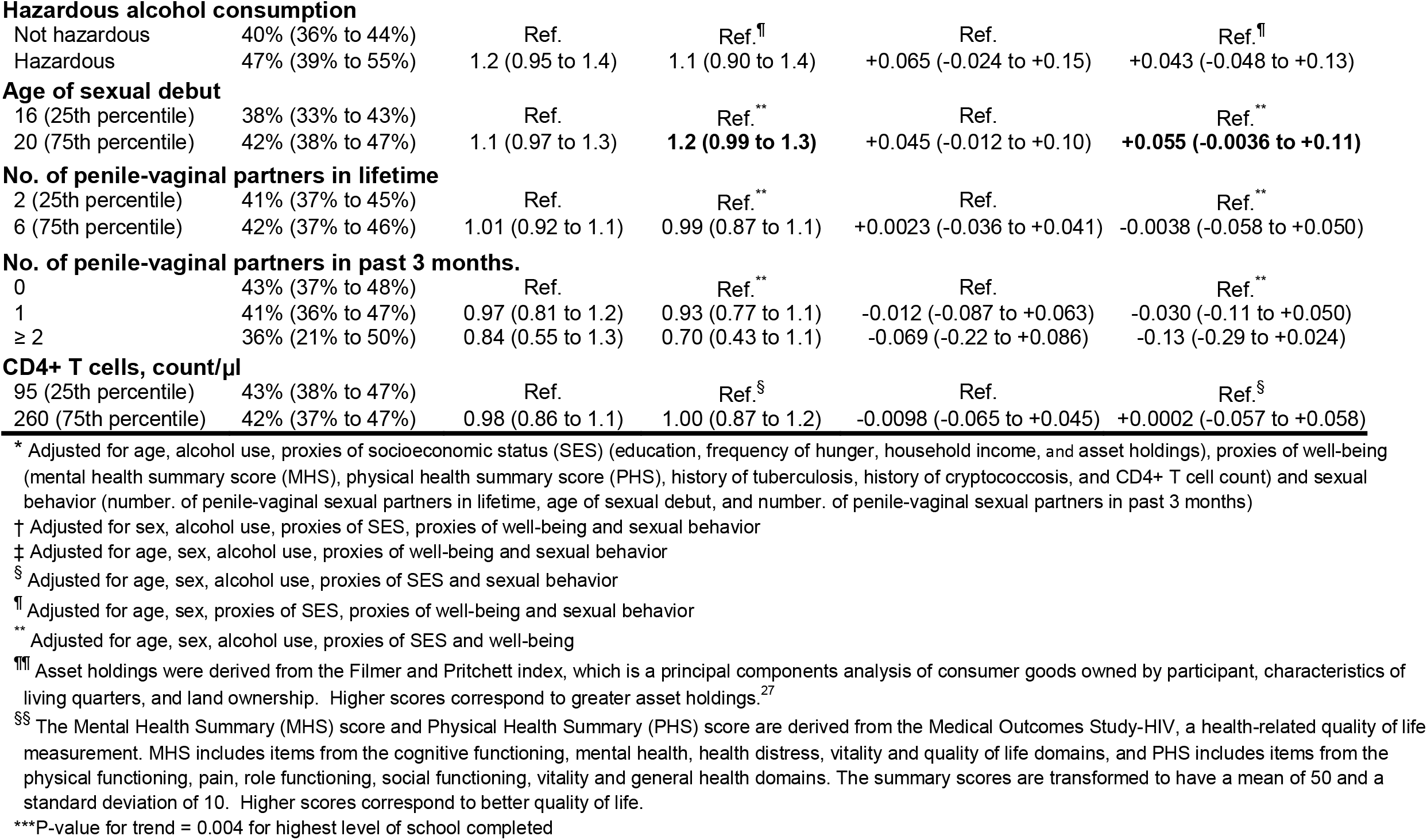
Relationship between a variety of participant characteristics (exposures) and KSHV antibody positivity (outcome) among HIV-infected adults in Uganda just prior to initiation of antiretroviral therapy.

| Characteristic | Percent KSHV<br>antibody positive<br>(95% CI) | Prevalence ratio |  | Prevalence difference |  |
| --- | --- | --- | --- | --- | --- |
|  |  | Unadjusted<br>(95% CI) | Adjusted<br>(95% CI) | Unadjusted<br>(95% CI) | Adjusted<br>(95% CI) |
| **Sex, biological** |  |  |  |  |  |
| Female | 36% (32% to 40%) | Ref. | Ref.* | Ref. | Ref.* |
| Male | 54% (48% to 61%) | **1.5 (1.3 to 1.8)** | **1.6 (1.3 to 1.9)** | **+0.18 (+0.11 to +0.26)** | **+0.20 (+0.11 to +0.30)** |
| **Age, years** |  |  |  |  |  |
| 28 (25th percentile) | 39% (35% to 44%) | Ref. | Ref.<sup>†</sup> | Ref. | Ref.<sup>†</sup> |
| 40 (75th percentile) | 43% (38% to 48%) | 1.1 (0.97 to 1.3) | 0.96 (0.85 to 1.1) | +0.040 (-0.0096 to +0.090) | -0.014 (-0.067 to +0.039) |
| **Highest level of school completed** |  |  |  |  |  |
| Secondary or tertiary | 33% (24% to 42%) | Ref. | Ref.<sup>‡ ***</sup> | Ref. | Ref.<sup>‡ ***</sup> |
| Some secondary | 39% (28% to 49%) | 1.2 (0.79 to 1.7) | 1.2 (0.82 to 1.8) | +0.056 (-0.084 to +0.20) | +0.065 (-0.068 to +0.20) |
| Primary | 41% (33% to 49%) | 1.3 (0.90 to 1.7) | 1.4 (0.97 to 1.9) | +0.081 (-0.037 to +0.20) | +0.11 (-0.0073 to +0.22) |
| Some primary | 45% (39% to 52%) | 1.4 (1.02 to 1.9) | 1.5 (1.1 to 2.09) | +0.12 (+0.016 to +0.23) | +0.16 (+0.054 to +0.26) |
| No school | 45% (35% to 55%) | 1.4 (0.96 to 2.0) | **1.6 (1.1 to 2.3)** | +0.12 (-0.016 to +0.26) | **+0.17 (+0.040 to +0.31)** |
| **Monthly household income, in US dollars** |  |  |  |  |  |
| 5.90 (25th percentile) | 44% (40% to 49%) | Ref. | Ref.<sup>‡</sup> | Ref. | Ref.<sup>‡</sup> |
| 41 (75th percentile) | 42% (37% to 47%) | 0.95 (0.84 to 1.06) | 0.93 (0.82 to 1.05) | -0.024 (-0.074 to +0.026) | -0.033 (-0.085 to +0.019) |
| **Asset holdings<sup>¶¶</sup>** |  |  |  |  |  |
| -1.5 (25th percentile) | 44% (40% to 49%) | Ref. | Ref.<sup>‡</sup> | Ref. | Ref.<sup>‡</sup> |
| 0.99 (75th percentile) | 40% (35% to 45%) | 0.95 (0.84 to 1.06) | 0.91 (0.79 to 1.05) | -0.040 (-0.099 to +0.019) | -0.042 (-0.10 to +0.018) |
| **Frequency of hunger in household** |  |  |  |  |  |
| Often | 48% (33% to 62%) | Ref. | Ref.<sup>‡</sup> | Ref. | Ref.<sup>‡</sup> |
| Sometimes | 43% (35% to 50%) | 0.90 (0.63 to 1.3) | 0.92 (0.66 to 1.3) | -0.049 (-0.21 to +0.11) | -0.037 (-0.20 to +0.12) |
| Seldom | 46% (37% to 55%) | 0.96 (0.67 to 1.4) | 0.93 (0.65 to 1.3) | -0.020 (-0.19 to +0.15) | -0.035 (-0.20 to +0.13) |
| Never | 39% (34% to 44%) | 0.83 (0.60 to 1.2) | 0.78 (0.56 to 1.1) | -0.082 (-0.23 to +0.071) | -0.11 (-0.26 to +0.046) |
| **Mental health summary score<sup>§§</sup>** |  |  |  |  |  |
| 47 (25th percentile) | 41% (36% to 46%) | Ref. | Ref.<sup>§</sup> | Ref. | Ref.<sup>§</sup> |
| 59 (75th percentile) | 42% (37% to 46%) | 1.02 (0.89 to 1.2) | 1.04 (0.90 to 1.2) | +0.0093 (-0.050 to +0.069) | +0.016 (-0.045 to +0.076) |
| **Physical health summary score<sup>§§</sup>** |  |  |  |  |  |
| 46 (25th percentile) | 37% (31% to 43%) | Ref. | Ref.<sup>§</sup> | Ref. | Ref.<sup>§</sup> |
| 60 (75th percentile) | 44% (39% to 49%) | 1.2 (0.98 to 1.5) | **1.3 (1.03 to 1.5)** | +0.071 (-0.0072 to +0.15) | **+0.094 (+0.015 to +0.17)** |
| **Tuberculosis, history of** |  |  |  |  |  |
| No | 42% (38% to 46%) | Ref. | Ref.<sup>§</sup> | Ref. | Ref.<sup>§</sup> |
| Yes | 38% (26% to 49%) | 0.89 (0.65 to 1.2) | 0.83 (0.60 to 1.2) | -0.045 (-0.17 to +0.076) | -0.071 (-0.19 to +0.048) |
| **Cryptococcosis, history of** |  |  |  |  |  |
| No | 42% (38% to 46%) | Ref. | Ref.<sup>§</sup> | Ref. | Ref.<sup>§</sup> |
| Yes | 28% (1.2% to 56%) | 0.68 (0.26 to 1.8) | 0.76 (0.31 to 1.9) | -0.13 (-0.41 to +0.14) | -0.098 (-0.39 to +0.19) |
| Hazardous alcohol consumption |  |  |  |  |  |
| Not hazardous | 40% (36% to 44%) | Ref. | Ref.<sup>††</sup> | Ref. | Ref.<sup>††</sup> |
| Hazardous | 47% (39% to 55%) | 1.2 (0.95 to 1.4) | 1.1 (0.90 to 1.4) | +0.065 (-0.024 to +0.15) | +0.043 (-0.048 to +0.13) |
| Age of sexual debut |  |  |  |  |  |
| 16 (25th percentile) | 38% (33% to 43%) | Ref. | Ref.<sup>**</sup> | Ref. | Ref.<sup>**</sup> |
| 20 (75th percentile) | 42% (38% to 47%) | 1.1 (0.97 to 1.3) | **1.2 (0.99 to 1.3)** | +0.045 (-0.012 to +0.10) | **+0.055 (-0.0036 to +0.11)** |
| No. of penile-vaginal partners in lifetime |  |  |  |  |  |
| 2 (25th percentile) | 41% (37% to 45%) | Ref. | Ref.<sup>**</sup> | Ref. | Ref.<sup>**</sup> |
| 6 (75th percentile) | 42% (37% to 46%) | 1.01 (0.92 to 1.1) | 0.99 (0.87 to 1.1) | +0.0023 (-0.036 to +0.041) | -0.0038 (-0.058 to +0.050) |
| No. of penile-vaginal partners in past 3 months. |  |  |  |  |  |
| 0 | 43% (37% to 48%) | Ref. | Ref.<sup>**</sup> | Ref. | Ref.<sup>**</sup> |
| 1 | 41% (36% to 47%) | 0.97 (0.81 to 1.2) | 0.93 (0.77 to 1.1) | -0.012 (-0.087 to +0.063) | -0.030 (-0.11 to +0.050) |
| ≥ 2 | 36% (21% to 50%) | 0.84 (0.55 to 1.3) | 0.70 (0.43 to 1.1) | -0.069 (-0.22 to +0.086) | -0.13 (-0.29 to +0.024) |
| CD4+ T cells, count/μl |  |  |  |  |  |
| 95 (25th percentile) | 43% (38% to 47%) | Ref. | Ref.<sup>§</sup> | Ref. | Ref.<sup>§</sup> |
| 260 (75th percentile) | 42% (37% to 47%) | 0.98 (0.86 to 1.1) | 1.00 (0.87 to 1.2) | -0.0098 (-0.065 to +0.045) | +0.0002 (-0.057 to +0.058) |

\* Adjusted for age, alcohol use, proxies of socioeconomic status (SES) (education, frequency of hunger, household income, and asset holdings), proxies of well-being (mental health summary score (MHS), physical health summary score (PHS), history of tuberculosis, history of cryptococcosis, and CD4+ T cell count) and sexual behavior (number. of penile-vaginal sexual partners in lifetime, age of sexual debut, and number. of penile-vaginal sexual partners in past 3 months)
† Adjusted for sex, alcohol use, proxies of SES, proxies of well-being and sexual behavior
‡ Adjusted for age, sex, alcohol use, proxies of well-being and sexual behavior
§ Adjusted for age, sex, alcohol use, proxies of SES and sexual behavior
†† Adjusted for age, sex, proxies of SES, proxies of well-being and sexual behavior
\*\* Adjusted for age, sex, alcohol use, proxies of SES and well-being
††† Asset holdings were derived from the Filmer and Pritchett index, which is a principal components analysis of consumer goods owned by participant, characteristics of living quarters, and land ownership. Higher scores correspond to greater asset holdings.<sup>27</sup>
§§ The Mental Health Summary (MHS) score and Physical Health Summary (PHS) score are derived from the Medical Outcomes Study-HIV, a health-related quality of life measurement. MHS includes items from the cognitive functioning, mental health, health distress, vitality and quality of life domains, and PHS includes items from the physical functioning, pain, role functioning, social functioning, vitality and general health domains. The summary scores are transformed to have a mean of 50 and a standard deviation of 10. Higher scores correspond to better quality of life.
\*\*\*P-value for trend = 0.004 for highest level of school completed

## Discussion

The advent of the HIV pandemic transformed KS from a rare medical curiosity to a multi-continent public health problem. Forty years later, despite successes in the control of HIV infection, persons living with HIV infection still represent the group at highest risk for KS. Many areas in sub-Saharan Africa have unfortunately lagged behind much of the world in achieving population-wide treatment of HIV infection, leaving the region amongst the world’s hotspots of KS.^47-50^ As such, to understand the threat of KS in Africa, it is relevant to focus on PLHIV. To address this, we measured the seroprevalence and determinants of the necessary causal agent of KS — KSHV — in a large and well-characterized group of adult PLHIV whose consecutive sampling in a municipal setting bolsters their representativeness of the larger target population. In a measurement domain that has been plagued by lack of agreement,^51-53^ we used a suite of time-tested KSHV antibody assays whose combination performance has been exemplary in available gold standard reference populations. We found a high prevalence of KSHV antibody positivity in this population — 42% — and applied various correction and sensitivity approaches to yield a range between 30% and 51%. We also found that biologic sex and education level attained had the largest-magnitude influence on KSHV seroprevalence.

Our finding of 42% KSHV antibody positivity in Uganda among adult PLHIV adds another estimate to an already heterogeneous lot. A systematic review of published literature from 7 East African countries (Djibouti, Eritrea, Ethiopia, Kenya, Somalia, Tanzania, and Uganda) reveals 9 other studies yielding a range of KSHV antibody positivity between 29% and 83% among adult PLHIV (Table 4). While not a probability sample of the source population, we believe that our sample of consecutive patients about to initiate ART at a prototypical municipal clinic is closely representative of the community population of adult PLHIV. Several of the prior studies feature equally convincing representative samples, including a door-to-door sample of households,^13, 14, 22, 54^ a probability sample of an entire country;^55^ and women attending antenatal clinic.^18, 20^ Other than geographic location, the main difference, in terms of important known determinants for KSHV infection, between prior studies and ours is the fraction of women. Accounting for sex, however, does not alter the heterogeneity. Among studies from East Africa in which the prevalence of KSHV antibody positivity in women could be isolated, values range from 36% and 70%. Of note, despite being just ∼160 kilometers (∼100 miles) apart and having nearly the same sex distribution, the prevalence of KSHV antibody positivity in our sample differed by an absolute -41% compared to Nalwoga et al.^20^

**Table 4.** Description of the study population, method of detection of antibodies to KSHV, and prevalence of KSHV antibody positivity among studies, including the present work, conducted in East African populations and identified in a systematic review of the published literature regarding KSHV antibody positivity among HIV-infected adults.

| Authors,<br>year of<br>publication | Country | Time<br>period | Study Sample | KSHV<br>antibody<br>testing<br>method | Percent<br>Female | No. of<br>HIV-infected<br>persons | Percent<br>(95%<br>confidence<br>interval)<br>KSHV-antibody-positive* |
| --- | --- | --- | --- | --- | --- | --- | --- |
| Wawer et al., 2000 | Uganda | 1999 to 2000 | Household residents, aged 15 to 59 years, living in one of 56 communities on secondary dirt roads in rural Rakai District. Of those approached, ~84% participated. | IFA targeting LANA and EIA targeting recombinant ORF65.2 | 54% | 76 | 29%<br>(19% to 40%) |
| Newton et al., 2003 | Uganda | 1994 to 1998 | Patients, ≥ 15 years of age, with a) non-malignant manifestations of HIV disease, recruited from outpatient unit of main municipal hospital in Kampala or b) new diagnosis of non-KS malignancies or benign tumors recruited from inpatient and outpatient units from several Kampala hospitals. | IFA targeting LANA | 66% | 422 | 41%<br>(36% to 46%) |
| Current study | Uganda | 2005 to 2008 and 2011 to 2013 | Patients, ≥ 18 years of age, newly initiating antiretroviral therapy at and residing within 60 kilometers of a municipal clinic in Mbarara | IFA targeting lytic antigens and EIAs, separately targeting synthetic K8.1 and ORF65 | 69% | 727 | 42%<br>(38% to 46%)<br>M: 54%<br>(47% to 61%)<br>F: 36%<br>(32% to 40%) |
| Butler et al., 2010 | Uganda | 2002 | All household residents, > 15 years of age, living in rural Buziika B Parish in Mukono District. Of those approached, 99% participated. | EIA targeting synthetic K8.1 | 61% | 81 | 44%<br>(33% to 56%)<br>M: 50%<br>(32% to 68%)<br>F: 40%<br>(27% to 56%) |
| Shebl et al., 2011 | Uganda | 2004 to 2005 | Persons, aged 15 to 59 years who had already experienced sexual debut, identified from a countrywide two-stage cluster sample of households. Of those approached, ~79% participated. | EIAs, separately targeting synthetic K8.1 and ORF65 | 55% | 138 | 57%<br>(48% to 65%)<br>M: 59%<br>(45% to 72%)<br>F: 55%<br>(44% to 66%) |
| Biryahwaho et al., 2010 | Uganda | 2004 to 2005 | Persons, aged 15 to 59 years, identified from a countrywide two-stage cluster sample of households. Of those approached, ~79% participated. | EIAs, separately targeting synthetic K8.1 and ORF65 | 53% | 81 | 57%<br>(48% to 65%)<br>M: 56%<br>(44% to 68%)<br>F: 57%<br>(46% to 69%) |
| Wakeham et al., 2011 | Uganda | 2003 to 2005 | Pregnant women, aged 14 years and older, identified at a government-funded antenatal clinic in Entebbe. Of those approached, ~90% participated. | EIAs, separately targeting recombinant K8.1 and ORF73 | 100% | 193 | 67%<br>(60% to 74%) |
| Lemma et al., 2009 | Ethiopia | 2006 to 2007 | Pregnant women, aged 15 years and older, identified at five regional antenatal clinics. | IFA targeting lytic antigens | 100% | 200 | 69%<br>(62% to 75%) |
| Nalwoga et al., 2015 | Uganda | 2003 to 2005 | Pregnant women, aged 14 years and older, identified at a government-funded antenatal clinic in Entebbe. Of those approached, ~70% participated. | EIAs, separately targeting recombinant K8.1 and ORF73 | 100% | 160 | 70%<br>(62% to 77%) |
| Nalwoga et al., 2018 | Uganda | 2014 to 2015 | All adults aged 18 to 103 years, residing in Kyamulibwa subcommunity of Kalungu District. Of those approached, ~95% participated. | EIAs, separately targeting recombinant K8.1 and ORF73 | 59% | 305 | 83%<br>(79% to 87%) |

\* In addition to percent of overall population who are KSHV antibody positive, separate estimates are shown, where available, for men (M) and women (F).

Aside from geographic differences across studies, the other difference is the method of antibody detection. First-generation assays for KSHV antibodies had a well-chronicled history of disagreement.^53^ Many of these assays have ceased being performed, but none of the surviving assays, which are all individual laboratory-derived “in house” protocols, have reached commercialization and accompanying high-fidelity standardization. Furthermore, there exists no study of agreement between the contemporary assays. Including the present study, seven different KSHV antibody detection platforms/algorithms are represented in the ten reports from East Africa to date (Table 4). The assay used by Wawer et al. featured an IFA targeted only at antibodies against KSHV latency-associated antigens (and a less sensitive lytic antigen); this may explain the low KSHV seroprevalence (29%) in this work.^22^ This explanation cannot, however, account for the difference between our work and all of the other community-based studies given that each study was nominally able to detect antibodies against lytic-phase KSHV antigens. Of note, our IFA, in addition to detecting antibodies to lytic-phase antigens, can also detect participants who solely have antibodies to latency-associated antigens. When we corrected our KSHV antibody positivity estimates to account for a range of assay sensitivity/specificity imperfection as low as 85% or altered our rule to allow reactivity on any single assay to count as overall positivity, our upper bound estimate still only reached 51%. Thus, the difference between our seroprevalence finding and the others is not readily explained by moderate differences in sensitivity or how we constructed our combined assay algorithm. In summary, the different antibody assays used across studies makes formal comparison difficult.

Biologic sex and level of education were the largest-magnitude determinants of KSHV antibody positivity. Higher prevalence of KSHV infection among men has been observed in several other studies from East Africa^13, 55-58^ but not all.^22, 54, 59^ The absolute difference we estimated (+20% among men), a value that is adjusted for many factors and represents a marginal measure of association, is the largest estimate of sex-derived differential for KSHV transmission of which we are aware in East Africa. The mechanism underlying the effect of male sex, which likely occurs in large part prior to puberty, is not understood but possibilities include sex-related differences in innate immune response^60, 61^ and behavioral exposures. A protective effect of higher education has also been observed in several other studies from East Africa^16, 18, 24, 55, 57^ but not all.^22, 62^ We speculate that education is a marker of socioeconomic status which influences some heretofore undocumented behavioral practice that transmits KSHV (e.g., some practice in which saliva is spread^63^). Regarding a role for sexual practices in the transmission of KSHV, the only association we found was a paradoxical higher KSHV seroprevalence with older age of sexual debut. We are unable to explain this low-magnitude association, but it fails to support sexual transmission of KSHV among adults. A lack of evidence for sexual transmission of KSHV has also been concluded in several other studies from this region.^13, 22, 24, 54^

Several limitations merit mention. First, the facility-based nature of our sampling precludes our ability to claim a pristinely representative sample of adult PLHIV. Specifically, we missed those who became HIV-infected but did not seek care, and, to the extent this is related to sex or education, our seroprevalence estimate may be biased. Second, regarding the measurement of KSHV antibodies, we earlier demonstrated how our antibody positivity estimate would be altered if assay sensitivity and specificity were as low as 85%. It is conceivable assay accuracy could even be lower but proving this will require a transformative finding in the field regarding assay gold standard construction. Finally, regarding confounding, residual confounding (particularly unmeasured) is a threat to some of our weak-magnitude analytic findings (e.g., older age at sexual debut). Notably, because sex is genetic in origin, there are few proximal causes, and, as depicted by our DAG, confounding of any relationships influenced by sex is highly unlikely.

There are several implications of our findings. The most prominent stems from the sizeable difference between our seroprevalence estimate (42%) and several others (up to 83%; Table 4) in the region. This disparity is explained either by differences in antibody assay performance or in the distribution of some very influential causative factor(s) for KSHV transmission. Resolving this will be critical and informative for the field. Inter-assay comparison studies using identical local specimens from East Africa are the requisite next step. Another implication is that, to the extent HIV infection does not influence KSHV acquisition (as most data indicate),^18, 21, 27, 54, 64^ our prevalence findings may, after standardization for sex distribution, also depict KSHV prevalence in the general population of East Africa. In turn, serial monitoring of KSHV seroprevalence in HIV-infected clinic populations could be an efficient way to monitor general population trends. In terms of mechanisms for KSHV transmission, our findings provide further evidence against any substantial magnitude of transmission in adulthood (including via sexual contact) in Africa. This is in contrast to the sexual transmission of KSHV that occurs among adults (men who have sex with men) in the U.S., Europe, and Australia.^65-70^ Instead of sexual transmission in adulthood, attention should be focused on KSHV transmission in childhood, particularly the mechanism underlying higher risk among boys. Finally, higher KSHV seroprevalence in men may explain, in part, the higher incidence of KS among men in Africa, especially HIV-uninfected men.^1, 71-73^

In summary, among adult PLHIV in western Uganda, we found that KSHV seroprevalence is ∼40%, with little change in estimate after various correction approaches accommodating uncertainty in antibody assay accuracy. Biological sex and education are the largest-magnitude determinants of KSHV infection; these findings motivate continued research regarding mechanism. The discrepancy between our KSHV seroprevalence and others in the region calls for formal inter-assay comparison studies using identical local East African specimens.

## Supporting information

Supplemental search strategy

## Data Availability

All data produced in the present study are available upon reasonable request to the authors.

## References

1. Cook-Mozaffari P, Newton R, Beral V, Burkitt DP. The geographical distribution of Kaposi’s sarcoma and of lymphomas in Africa before the AIDS epidemic. British Journal of Cancer. 78(11):1521–8, 1998.

2. Chaabna K, Bray F, Wabinga HR, Chokunonga E, Borok M, Vanhems P, Forman D, Soerjomataram I. Kaposi sarcoma trends in Uganda and Zimbabwe: a sustained decline in incidence? International Journal of Cancer. 133(5):1197–203, 2013.

3. Parkin DM, Sitas F, Chirenje M, Stein L, Abratt R, Wabinga H. Part I: cancer in Indigenous Africans-burden, distribution, and trends. The Lancet Oncology. 9(7):683–92, 2008.

4. Wabinga HR, Parkin DM, Wabwire-Mangen F, Mugerwa JW. Cancer in Kampala, Uganda, in 1989-91: changes in incidence in the era of AIDS. International Journal of Cancer. 54(1):26–36, 1993.

5. Liu Z, Fang Q, Zuo J, Minhas V, Wood C, Zhang T. The world-wide incidence of Kaposi’s sarcoma in the HIV/AIDS era. HIV Medicine. 19(5):355–64, 2018.

6. Martin J, Wenger M, Busakhala N, Buziba N, Bwana M, Muyindike W, Mbabazi R, Amerson E, Yiannoutsos C, Musick B, LeBoit P, McCalmont T, Ruben B, Maurer T, Wools-Kaloustian K. Prospective evaluation of the impact of potent antiretroviral therapy on the incidence of Kaposi’s sarcoma in East Africa: findings from the International Epidemiologic Databases to Evaluate AIDS (IeDEA) consortium. Infectious Agents and Cancer. 7(1):O19, 2012.

7. Beral V, Peterman TA, Berkelman RL, Jaffe HW. Kaposi’s sarcoma among persons with AIDS: a sexually transmitted infection? The Lancet. 335(8682):123–8, 1990.

8. Chang Y, Cesarman E, Pessin MS, Lee F, Culpepper J, Knowles DM, Moore PS. Identification of herpesvirus-like DNA sequences in AIDS-associated Kaposi’s sarcoma. Science. 266(5192):1865–9, 1994.

9. Mesri EA, Feitelson MA, Munger K. Human viral oncogenesis: a cancer hallmarks analysis. Cell Host & Microbe. 15(3):266–82, 2014.

10. da Silva SR, de Oliveira DE. HIV, EBV and KSHV: viral cooperation in the pathogenesis of human malignancies. Cancer Letters. 305(2):175–85, 2011.

11. La Ferla L, Pinzone MR, Nunnari G, Martellotta F, Lleshi A, Tirelli U, De Paoli P, Berretta M, Cacopardo B. Kaposi’ s sarcoma in HIV-positive patients: the state of art in the HAART-era. European Review for Medical and Pharmacological Sciences. 17(17):2354–65, 2013.

12. Cancer incidence in five continents volume XI (electronic version). Lyon: International agency for research on cancer (IARC); 2017; Available from: http://ci5.iarc.fr. Accessed 2019.

13. Butler LM, Were WA, Balinandi S, Downing R, Dollard S, Neilands TB, Gupta S, Rutherford GW, Mermin J. Human herpesvirus 8 infection in children and adults in a population-based study in rural Uganda. The Journal of Infectious Diseases. 203(5):625–34, 2011.

14. Nalwoga A, Cose S, Nash S, Miley W, Asiki G, Kusemererwa S, Yarchoan R, Labo N, Whitby D, Newton R. Relationship between anemia, malaria coinfection, and Kaposi sarcoma-associated herpesvirus seropositivity in a population-based study in rural Uganda. The Journal of Infectious Diseases. 218(7):1061–5, 2018.

15. Marcelin AG, Grandadam M, Flandre P, Nicand E, Milliancourt C, Koeck JL, Philippon M, Teyssou R, Agut H, Dupin N, Calvez V. Kaposi’s sarcoma herpesvirus and HIV-1 seroprevalences in prostitutes in Djibouti. Journal of Medical Virology. 68(2):164–7, 2002.

16. Lavreys L, Chohan B, Ashley R, Richardson BA, Corey L, Mandaliya K, Ndinya-Achola JO, Kreiss JK. Human herpesvirus 8: seroprevalence and correlates in prostitutes in Mombasa, Kenya. The Journal of Infectious Diseases. 187(3):359–63, 2003.

17. Lemma E, Constantine NT, Kassa D, Messele T, Mindaye T, Taye G, Abebe A, Tamene W, Tebje M, Gebremeskel W, Adane A, Gezahegn N. Human herpesvirus 8 infection in HIV-1-infected and uninfected pregnant women in Ethiopia. Ethiopian Medical Journal. 47(3):205–11, 2009.

18. Wakeham K, Webb EL, Sebina I, Muhangi L, Miley W, Johnson WT, Ndibazza J, Elliott AM, Whitby D, Newton R. Parasite infection is associated with Kaposi’s sarcoma associated herpesvirus (KSHV) in Ugandan women. Infectious Agents and Cancer. 6(1):15, 2011.

19. Butler LM, Dorsey G, Hladik W, Rosenthal PJ, Brander C, Neilands TB, Mbisa G, Whitby D, Kiepiela P, Mosam A, Mzolo S, Dollard SC, Martin JN. Kaposi sarcoma-associated herpesvirus (KSHV) seroprevalence in population-based samples of African children: evidence for at least 2 patterns of KSHV transmission. The Journal of Infectious Diseases. 200(3):430–8, 2009.

20. Nalwoga A, Cose S, Wakeham K, Miley W, Ndibazza J, Drakeley C, Elliott A, Whitby D, Newton R. Association between malaria exposure and Kaposi’s sarcoma-associated herpes virus seropositivity in Uganda. Tropical Medicine & International Health. 20(5):665–72, 2015.

21. Newton R, Ziegler J, Bourboulia D, Casabonne D, Beral V, Mbidde E, Carpenter L, Parkin DM, Wabinga H, Mbulaiteye S, Jaffe H, Weiss R, Boshoff C. Infection with Kaposi’s sarcoma-associated herpesvirus (KSHV) and human immunodeficiency virus (HIV) in relation to the risk and clinical presentation of Kaposi’s sarcoma in Uganda. British Journal of Cancer. 89(3):502–4, 2003.

22. Wawer MJ, Eng SM, Serwadda D, Sewankambo NK, Kiwanuka N, Li C, Gray RH. Prevalence of Kaposi sarcoma-associated herpesvirus compared with selected sexually transmitted diseases in adolescents and young adults in rural Rakai district, Uganda. Sexually Transmitted Diseases. 28(2):77–81, 2001.

23. Maskew M, Macphail AP, Whitby D, Egger M, Wallis CL, Fox MP. Prevalence and predictors of Kaposi sarcoma herpesvirus seropositivity: a cross-sectional analysis of HIV-infected adults initiating ART in Johannesburg, South Africa. Infectious Agents and Cancer. 6:22, 2011.

24. Klaskala W, Brayfield BP, Kankasa C, Bhat G, West JT, Mitchell CD, Wood C. Epidemiological characteristics of human herpesvirus-8 infection in a large population of antenatal women in Zambia. Journal of Medical Virology. 75(1):93–100, 2005.

25. Nzivo MM, Lwembe RM, Odari EO, Kang’ethe JM, Budambula NLM. Prevalence and risk factors of Human Herpes Virus type 8 (HHV-8), Human Immunodeficiency Virus-1 (HIV-1), and syphilis among female sex workers in Malindi, Kenya. Interdisciplinary Perspectives on Infectious Diseases. 2019:5345161, 2019.

26. Enbom M, Tolfvenstam T, Ghebrekidan H, Ruden U, Grandien M, Wahren B, Linde A. Seroprevalence of human herpes virus 8 in different Eritrean population groups. Journal of Clinical Virology. 14(3):167–72, 1999.

27. Filmer D, Pritchett LH. Estimating wealth effects without expenditure data--or tears: an application to educational enrollments in states of India. Demography. 38(1):115–32, 2001.

28. Stangl AL, Wamai N, Mermin J, Awor AC, Bunnell RE. Trends and predictors of quality of life among HIV-infected adults taking highly active antiretroviral therapy in rural Uganda. AIDS Care. 19(5):626–36, 2007.

29. Saunders JB, Aasland OG, Babor TF, de la Fuente JR, Grant M. Development of the Alcohol Use Disorders Identification Test (AUDIT): WHO collaborative project on early detection of persons with harmful alcohol consumption--II. Addiction. 88(6):791–804, 1993.

30. Pau CP, Lam LL, Spira TJ, Black JB, Stewart JA, Pellett PE, Respess RA. Mapping and serodiagnostic application of a dominant epitope within the human herpesvirus 8 ORF 65-encoded protein. Journal of Clinical Microbiology. 36(6):1574–7, 1998.

31. Spira TJ, Lam L, Dollard SC, Meng YX, Pau CP, Black JB, Burns D, Cooper B, Hamid M, Huong J, Kite-Powell K, Pellett PE. Comparison of serologic assays and PCR for diagnosis of human herpesvirus 8 infection. Journal of Clinical Microbiology. 38(6):2174–80, 2000.

32. Dollard SC, Nelson KE, Ness PM, Stambolis V, Kuehnert MJ, Pellett PE, Cannon MJ. Possible transmission of human herpesvirus-8 by blood transfusion in a historical United States cohort. Transfusion. 45(4):500–3, 2005.

33. Lennette ET, Blackbourn DJ, Levy JA. Antibodies to human herpesvirus type 8 in the general population and in Kaposi’s sarcoma patients. Lancet. 348(9031):858–61, 1996.

34. Dollard SC, Butler LM, Jones AMG, Mermin JH, Chidzonga M, Chipato T, Shiboski CH, Brander C, Mosam A, Kiepiela P, Hladik W, Martin JN. Substantial regional differences in human herpesvirus 8 seroprevalence in sub-Saharan Africa: Insights on the origin of the “Kaposi’s sarcoma belt”. International Journal of Cancer. 127(10):2395–401, 2010.

35. Pellett PE, Wright DJ, Engels EA, Ablashi DV, Dollard SC, Forghani B, Glynn SA, Goedert JJ, Jenkins FJ, Lee TH, Neipel F, Todd DS, Whitby D, Nemo GJ, Busch MP. Multicenter comparison of serologic assays and estimation of human herpesvirus 8 seroprevalence among US blood donors. Transfusion. 43(9):1260–8, 2003.

36. Cannon MJ, Operskalski EA, Mosley JW, Radford K, Dollard SC. Lack of evidence for human herpesvirus-8 transmission via blood transfusion in a historical US cohort. The Journal of Infectious Diseases. 199(11):1592–8, 2009.

37. Rogan WJ, Gladen B. Estimating prevalence from the results of a screening test. American Journal of Epidemiology. 107(1):71–6, 1978.

38. Lang Z, Reiczigel J. Confidence limits for prevalence of disease adjusted for estimated sensitivity and specificity. Preventive Veterinary Medicine. 113(1):13–22, 2014.

39. Hernán MA, Hernández-Díaz S, Werler MM, Mitchell AA. Causal knowledge as a prerequisite for confounding evaluation: an application to birth defects epidemiology. American Journal of Epidemiology. 155(2):176–84, 2002.

40. Shrier I, Platt RW. Reducing bias through directed acyclic graphs. BMC Medical Research Methodology. 8(1):70, 2008.

41. Westreich D, Greenland S. The table 2 fallacy: presenting and interpreting confounder and modifier coefficients. American Journal of Epidemiology. 177(4):292–8, 2013.

42. Cummings P. Estimating adjusted risk ratios for matched and unmatched data: An update. Stata Journal. 11(2):290–8, 2011.

43. Snowden JM, Rose S, Mortimer KM. Implementation of G-computation on a simulated data set: demonstration of a causal inference technique. American Journal of Epidemiology. 173(7):731–8, 2011.

44. Stata multivariate statistics reference manual : release 10: College Station, Tex. : StataCorp LP, [2007] ©2007; 2007.

45. Hosmer DW, Lemesbow S. Goodness of fit tests for the multiple logistic regression model. Communications in Statistics - Theory and Methods. 9(10):1043–69, 1980.

46. Cesarman E, Damania B, Krown SE, Martin J, Bower M, Whitby D. Kaposi sarcoma. Nature Reviews Disease Primers. 5(1):9, 2019.

47. Chinula L, Moses A, Gopal S. HIV-associated malignancies in sub-Saharan Africa: progress, challenges, and opportunities. Current Opinion in HIV and AIDS. 12(1):89–95, 2017.

48. Haq H, Elyanu P, Bulsara S, Bacha JM, Campbell LR, El-Mallawany NK, Keating EM, Kisitu GP, Mehta PS, Rees CA, Slone JS, Kekitiinwa AR, Matshaba M, Mizwa MB, Mwita L, Schutze GE, Wanless SR, Scheurer ME, Lubega J. Association between antiretroviral therapy and cancers among children living with HIV in sub-saharan Africa. Cancers (Basel). 13(6), 2021.

49. Lin LL, Lakomy DS, Chiao EY, Strother RM, Wirth M, Cesarman E, Borok M, Busakhala N, Chibwesha CJ, Chinula L, Ndlovu N, Orem J, Phipps W, Sewram V, Vogt SL, Sparano JA, Mitsuyasu RT, Krown SE, Gopal S. Clinical trials for treatment and prevention of HIV-associated malignancies in sub-saharan Africa: building capacity and overcoming barriers. JCO Global Oncology. 6:1134–46, 2020.

50. Nascimento MC, de Souza VA, Sumita LM, Freire W, Munoz F, Kim J, Pannuti CS, Mayaud P. Comparative study of Kaposi’s sarcoma-associated herpesvirus serological assays using clinically and serologically defined reference standards and latent class analysis. Journal of Clinical Microbiology. 45(3):715–20, 2007.

51. Schatz O, Monini P, Bugarini R, Neipel F, Schulz TF, Andreoni M, Erb P, Eggers M, Haas J, Buttò S, Lukwiya M, Bogner JR, Yaguboglu S, Sheldon J, Sarmati L, Goebel FD, Hintermaier R, Enders G, Regamey N, Wernli M, Stürzl M, Rezza G, Ensoli B. Kaposi’s sarcoma-associated herpesvirus serology in Europe and Uganda: multicentre study with multiple and novel assays. Journal of Medical Virology. 65(1):123–32, 2001.

52. Rabkin CS, Schulz TF, Whitby D, Lennette ET, Magpantay LI, Chatlynne L, Biggar RJ. Interassay correlation of human herpesvirus 8 serologic tests. HHV-8 interlaboratory collaborative group. The Journal of Infectious Diseases. 178(2):304–9, 1998.

53. Shebl FM, Dollard SC, Pfeiffer RM, Biryahwaho B, Amin MM, Munuo SS, Hladik W, Parsons R, Graubard BI, Mbulaiteye SM. Human herpesvirus 8 seropositivity among sexually active adults in Uganda. PLoS One. 6(6):e21286, 2011.

54. Biryahwaho B, Dollard SC, Pfeiffer RM, Shebl FM, Munuo S, Amin MM, Hladik W, Parsons R, Mbulaiteye SM. Sex and geographic patterns of human herpesvirus 8 infection in a nationally representative population-based sample in Uganda. The Journal of Infectious Diseases. 202(9):1347–53, 2010.

55. Nalwoga A, Webb EL, Chihota B, Miley W, Walusimbi B, Nassuuna J, Sanya RE, Nkurunungi G, Labo N, Elliott AM, Cose S, Whitby D, Newton R. Kaposi’s sarcoma-associated herpesvirus seropositivity is associated with parasite infections in Ugandan fishing communities on Lake Victoria islands. PLOS Neglected Tropical Diseases. 13(10):e0007776, 2019.

56. Caterino-de-Araujo A, Manuel RC, Del Bianco R, Santos-Fortuna E, Magri MC, Silva JM, Bastos R. Seroprevalence of human herpesvirus 8 infection in individuals from health care centers in Mozambique: potential for endemic and epidemic Kaposi’s sarcoma. Journal of Medical Virology. 82(7):1216–23, 2010.

57. Newton R, Labo N, Wakeham K, Miley W, Asiki G, Johnston WT, Whitby D. Kaposi sarcoma-associated herpesvirus in a rural Ugandan cohort, 1992-2008. The Journal of Infectious Diseases. 217(2):263–9, 2018.

58. DeSantis SM, Pau CP, Archibald LK, Nwanyanwu OC, Kazembe PN, Dobbie H, Jarvis WR, Jason J. Demographic and immune correlates of human herpesvirus 8 seropositivity in Malawi, Africa. International Journal of Infectious Diseases. 6(4):266–71, 2002.

59. Wang X, Zou Z, Deng Z, Liang D, Zhou X, Sun R, Lan K. Male hormones activate EphA2 to facilitate Kaposi’s sarcoma-associated herpesvirus infection: Implications for gender disparity in Kaposi’s sarcoma. PLOS Pathogens. 13(9):e1006580, 2017.

60. Nalwoga A, Nakibuule M, Marshall V, Miley W, Labo N, Cose S, Whitby D, Newton R. Risk factors for Kaposi’s sarcoma-associated herpesvirus DNA in blood and in saliva in rural Uganda. Clinical Infectious Diseases. 71(4):1055–62, 2020.

61. Brayfield BP, Phiri S, Kankasa C, Muyanga J, Mantina H, Kwenda G, West JT, Bhat G, Marx DB, Klaskala W, Mitchell CD, Wood C. Postnatal human herpesvirus 8 and human immunodeficiency virus type 1 infection in mothers and infants from Zambia. The Journal of Infectious Diseases. 187(4):559–68, 2003.

62. Butler LM, Neilands TB, Mosam A, Mzolo S, Martin JN. A population-based study of how children are exposed to saliva in KwaZulu-Natal Province, South Africa: implications for the spread of saliva-borne pathogens to children. Tropical Medicine & International Health. 15(4):442–53, 2010.

63. Phipps W, Saracino M, Selke S, Huang ML, Jaoko W, Mandaliya K, Wald A, Casper C, McClelland RS. Oral HHV-8 replication among women in Mombasa, Kenya. Journal of Medical Virology. 86(10):1759–65, 2014.

64. Martin JN, Ganem DE, Osmond DH, Page-Shafer KA, Macrae D, Kedes DH. Sexual transmission and the natural history of human herpesvirus 8 infection. The New England Journal of Medicine. 338(14):948–54, 1998.

65. Smith NA, Sabin CA, Gopal R, Bourboulia D, Labbet W, Boshoff C, Barlow D, Band B, Peters BS, de Ruiter A, Brown DW, Weiss RA, Best JM, Whitby D. Serologic evidence of human herpesvirus 8 transmission by homosexual but not heterosexual sex. The Journal of Infectious Diseases. 180(3):600–6, 1999.

66. Grulich AE, Cunningham P, Munier ML, Prestage G, Amin J, Ringland C, Whitby D, Kippax S, Kaldor JM, Rawlinson W. Sexual behaviour and human herpesvirus 8 infection in homosexual men in Australia. Sex Health. 2(1):13–8, 2005.

67. Martró E, Esteve A, Schulz TF, Sheldon J, Gambús G, Muñoz R, Whitby D, Casabona J. Risk factors for human Herpesvirus 8 infection and AIDS-associated Kaposi’s sarcoma among men who have sex with men in a European multicentre study. International Journal of Cancer. 120(5):1129–35, 2007.

68. Engels EA, Atkinson JO, Graubard BI, McQuillan GM, Gamache C, Mbisa G, Cohn S, Whitby D, Goedert JJ. Risk factors for human herpesvirus 8 infection among adults in the United States and evidence for sexual transmission. The Journal of Infectious Diseases. 196(2):199–207, 2007.

69. Osmond DH, Buchbinder S, Cheng A, Graves A, Vittinghoff E, Cossen CK, Forghani B, Martin JN. Prevalence of Kaposi sarcoma–associated herpesvirus infection in homosexual men at beginning of and during the HIV epidemic. JAMA. 287(2):221–5, 2002.

70. Melbye M, Cook PM, Hjalgrim H, Begtrup K, Simpson GR, Biggar RJ, Ebbesen P, Schulz TF. Risk factors for Kaposi’s-sarcoma-associated herpesvirus (KSHV/HHV-8) seropositivity in a cohort of homosexual men, 1981–1996. International Journal of Cancer. 77(4):543–8, 1998.

71. Chaabna K, Boniol M, de Vuyst H, Vanhems P, de Ávila Vitoria MA, Curado M-P. Geographical patterns of Kaposi’s sarcoma, nonHodgkin lymphomas, and cervical cancer associated with HIV infection in five African populations. European Journal of Cancer Prevention. 21(1):1–9, 2012.

72. Dedicoat M, Newton R. Review of the distribution of Kaposi’s sarcoma-associated herpesvirus (KSHV) in Africa in relation to the incidence of Kaposi’s sarcoma. British Journal of Cancer. 88(1):1–3, 2003.

