## Supplemental search strategy for "Prevalence and determinants of Kaposi’s sarcoma-associated herpesvirus (KSHV) antibody positivity among adults living with HIV in East Africa"

PubMed search strategy

**((((prevalen* OR inciden* OR epidemiolog* OR seroprevalen* OR sero-prevalen* OR seroepidemiolog* OR sero-epidemiolog* OR seropositiv* OR sero-positiv* OR seroepidemiologic studies[MeSH] OR prevalence[MeSH] OR incidence[MeSH]))** AND (("Africa, Eastern"[Mesh] OR East Africa OR Eastern Africa OR East African OR Eastern African OR Djibouti OR Eritrea OR Ethiopia OR Kenya OR Somalia OR Tanzania OR Uganda))) AND (("hiv infections"[MeSH Terms] OR ("hiv"[All Fields] AND ("infections"[All Fields] OR infection OR infected)))) AND (("herpesvirus 8, human"[MeSH Terms] OR "human herpesvirus 8"[All Fields] OR "hhv 8"[All Fields] OR "kshv"[All Fields] OR kaposi sarcoma herpesvirus))

Embase search strategy

**((prevalen* OR inciden* OR epidemiolog* OR seroprevalen* OR 'sero prevalen*' OR seroepidemiolog* OR 'sero epidemiolog*' OR seropositiv* OR 'sero positiv*' OR 'seroepidemiologic studies'/exp OR 'seroepidemiologic studies' OR (seroepidemiologic AND ('studies'/exp OR studies)) OR 'prevalence'/exp OR prevalence OR 'incidence'/exp OR incidence)** AND (('east africa'/exp OR 'east africa' OR (east AND ('africa'/exp OR africa)) OR 'eastern africa'/exp OR 'eastern africa' OR (eastern AND ('africa'/exp OR africa)) OR 'east african'/exp OR 'east african' OR (east AND ('african'/exp OR african)) OR 'eastern african'/exp OR 'eastern african' OR (eastern AND ('african'/exp OR african)) OR 'djibouti'/exp OR djibouti OR 'eritrea'/exp OR eritrea OR 'ethiopia'/exp OR ethiopia OR 'kenya'/exp OR kenya OR 'somalia'/exp OR somalia OR 'tanzania'/exp OR tanzania OR 'uganda'/exp OR uganda) AND ('human herpesvirus 8'/exp OR 'human herpesvirus 8' OR 'hhv 8' OR 'kshv' OR 'kaposi sarcoma herpesvirus'/exp OR 'kaposi sarcoma herpesvirus') AND (('hiv'/exp OR 'hiv') AND ('infections'/exp OR 'infections') OR 'hiv infections'/exp OR 'hiv infections' OR 'hiv infection'/exp OR 'hiv infection'))) AND ('article'/it OR 'article in press'/it)
